# Antemortem α-Synuclein levels from neuron-derived extracellular vesicles identify Lewy Body pathology in Parkinson’s Disease and Alzheimer Disease

**DOI:** 10.64898/2026.08.03.26359636

**Authors:** Tamil Iniyan Gunasekaran, Dolly Reyes-Dumeyer, Yian Gu, Olga Volpert, Doug Beauregard, Lammert Albers, Andrew Teich, Lawrence S. Honig, Richard Mayeux, Erez Eitan, Badri N. Vardarajan

## Abstract

**Importance:** Alzheimer disease (AD) is frequently accompanied by co-pathologies such as Lewy bodies, intracellular protein aggregates consisting of misfolded α-synuclein, ubiquitin and several other proteins. These aggregated proteins contribute to the pathological and clinical heterogeneity of AD and are associated with a more rapid progression. Cerebrospinal fluid (CSF) measurement of aggregated α-synuclein or skin biopsy histochemistry are the current biomarkers for Lewy body pathology in individuals with Parkinson’s disease and related synucleinopathies but are invasive, not easily scalable for large studies or practical in clinical practice. Neuron-derived extracellular vesicles (nEVs) provide a feasible and tolerable alternative to detect Lewy body pathology. nEVs exit across the blood-brain barrier and deliver neuron-derived proteins into the systemic circulation, offering direct access to brain-level protein concentrations from peripheral blood, not afforded by conventional plasma biomarkers.

**Objective:** To use nEVs extracted from plasma to measure brain-derived α-synuclein levels as a clinical biomarker of Lewy body pathology and synucleinopathy across clinically and pathologically characterized cohorts.

**Design, Setting, and Participants:** This multicohort observational study evaluated plasma nEV-derived α-synuclein levels in 1,304 individuals, including 794 (61%) individuals without dementia, 469 (36%) individuals with AD, and 41 (3%) individuals with PD across four cohorts. Postmortem validation was performed from autopsy data in 127 individuals, including 96 (76%) individuals without, and 31 (24%) with, Lewy body pathology. Cerebrospinal fluid (CSF) α-synuclein seed amplification assay data were available in a small subset of 54 (4.1%) of the group. Amyloid positron emission tomography (PET) data were available in 901(69.1%) of the total group.

**Exposure:** Clinical diagnosis, AD biomarker positivity, Lewy body pathology, CSF α-synuclein seed assay status, and amyloid PET positivity.

**Main Outcomes and Measures:** Plasma nEV-derived α-synuclein levels normalized to CD9 and assessed in relation to clinical diagnosis, AD biomarker status, neuropathologically confirmed Lewy body pathology, CSF seed assay results, and amyloid PET positivity.

**Results:** Compared with controls, plasma α-synuclein levels from nEVs were significantly elevated among individuals with PD (controls mean 0.51, SD=0.24 vs PD mean=0.69, SD=0.23, *P*=8.66×10^-^ □). nEV-derived α-synuclein levels were also significantly higher in P-tau181 and P- tau217 positive individuals with and without cognitive impairment compared to P-tau negative individuals. Elevated nEV-derived α-synuclein levels were subsequently validated in individuals with postmortem Lewy body pathology (28.6% higher mean levels, *P*=0.02). Among 54 individuals with CSF α-synuclein seed amplification assay, 8 (15%) were positive and showed a 18.2% increase in mean nEV α-synuclein levels compared to individuals with negative α- synuclein seed amplification. nEV-derived α-synuclein levels were also significantly elevated in amyloid PET positive individuals with and without dementia (5.8% higher mean levels, *P*=8.50E-03) and clinical AD (9.3% higher mean levels, *P*=6.38E-12).

**Conclusions and Relevance:** Plasma α-synuclein levels from nEVs likely reflect underlying synuclein pathology in the brain and have potential as a blood-based biomarker for detecting Lewy body pathology in PD and in AD and related dementias. nEVs cross the blood–brain barrier and carry neuron-derived cargo directly into the bloodstream, thus providing a unique window into brain α-synuclein levels that is not accessible through conventional plasma biomarkers.

## Introduction

Alpha-synuclein (α-synuclein) aggregation is a central pathological feature of a spectrum of neurodegenerative disorders collectively referred to as synucleinopathies, including Parkinson’s disease (PD), dementia with Lewy bodies (DLB), and Alzheimer’s disease (AD) with co-existing Lewy body pathology^1–3^. The presence of α-synuclein pathology is strongly associated with cognitive decline, neuropsychiatric symptoms, and rapid disease progression, underscoring the importance of detecting and quantifying α-synuclein in living patients^1,4,5^. However, despite its clinical relevance, reliable and scalable biomarkers for α-synuclein pathology remain limited^4,6^. Definitive diagnosis has traditionally relied on post-mortem neuropathological confirmation, while in-vivo assessments depend heavily on clinical criteria and neuroimaging, both of which have well-documented limitations in sensitivity and specificity, particularly at early disease stages^7,8^.

The potential of fluid-based biomarkers has transformed the landscape of neurodegenerative disease diagnosis^9^. Cerebrospinal fluid (CSF) α-synuclein seed amplification assays (SAA) is considered an accurate indicator of synuclein pathology, demonstrating high specificity and reasonable sensitivity for cortical Lewy body disease^10–12^. Skin biopsy^13,14^ has also been used to detect phosphorylated α-synuclein in individuals with PD, DLB, and multiple system atrophy. However, CSF collection and skin biopsy are invasive, limiting scalability, and are poorly suited for population-level screening or longitudinal monitoring^15^. Blood-based biomarkers offer a compelling alternative, but despite excellent performance of phosphorylated tau, (P-tau), neurofilament light chain (NfL), and others, detecting brain-derived pathological synuclein in plasma presents has presented formidable technical obstacles^16^. α-synuclein, for example, is expressed by erythrocytes and other non-neuronal cell types, making blood analysis of its level non-specific^17,18^.

An approach to overcome this challenge is the selective isolation of neuron-derived extracellular vesicles (nEVs) from peripheral blood. nEVs circulate in plasma and carry cargo including proteins and RNA that reflect the molecular state of neurons in the central nervous system^19,20^. By enriching for this neuronal fraction, it becomes possible to measure disease-relevant proteins such as α-synuclein by filtering the non-relevant α-synuclein coming from erythrocytes and other cell types^18,21–23^. Immunoaffinity-based platforms designed for nEVs isolation have demonstrated the ability to capture EVs expressing neuronal surface markers, providing a window into CNS pathology via a minimally invasive blood draw^19,20^.

Here, we report automated methods for nEVs enrichment (ExoSORT) followed by measurement and analysis of α-synuclein levels across several independent cohorts: The Estudio Familiar de Influencia Genetica en Alzheimer (EFIGA), the Washington Heights, Inwood, Columbia Aging Project (WHICAP), Bio-Hermes Study, and the Parkinson’s Progression Markers Initiative (PPMI)^24^. We further examined concordance with established gold standards, including post-mortem neuropathological confirmation and CSF seed amplification assay, and discuss the potential role of nEVs-derived α−synuclein as a component of a multi-marker diagnostic framework for neurodegenerative disease.

## Results

### Multi-cohort demographics

A total of 1,304 participants across EFIGA, WHICAP, Bio-Hermes, and PPMI, cohorts were included in this study (Table 1). Among all individuals, 61% were without dementia, 36.3% had AD or related dementia, and 3.1% had PD.

**Table 1.** Demographic characteristics across study cohorts Parkinson’s Precision Markers Initiative (PPMI)

| **Parkinson's Precision Markers Initiative (PPMI)** |  |  |  |
| --- | --- | --- | --- |
| **Characteristic** | **Healthy Controls (n=42)** | **Parkinson's disease (n=41)** | **P-value** |
| Women % | 24 (57%) | 22 (53%) | 0.75 |
| **nEV biomarkers, Mean (SD)** |  |  |  |
| α-Synuclein | 0.51 (24%) | 0.69 (23%) | <0.001 |
| **Columbia University EFIGA** |  |  |  |
| **Characteristic** | **Cognitively normal (n=29)** | **AD dementia (n=25)** | **P-value** |
| Women % | 17 (59%) | 18 (72%) | 0.18 |
| APOE ε4% | 38% | 60% | 0.90 |
| Age, Mean (SD) | 69.82 (7.35) | 68.17 (8.31) | 0.92 |
| CSF seed assay positive (%) | 2 (0.07) | 12 (0.48) | 0.002 |
| P-tau181 positive (%) | 15 (0.48) | 13 (0.48) | 0.98 |
| **nEV biomarkers, Mean (SD)** |  |  |  |
| α-Synuclein | 0.46 (0.18) | 0.52 (0.26) | 0.15 |
| **Columbia University WHICAP** |  |  |  |
| **Characteristic** | **Cognitively normal (n=120)** | **AD dementia (n=142)** | **P-value** |
| Women % | 73 (61%) | 92 (65%) | 0.51 |
| APOE ε4% | 31% | 37% | 0.45 |
| Age, Mean (SD) | 84.31 (7.35) | 87.1 (6.06) | <0.001 |
| P-tau181 positive (%) | 47 (42%) | 42 (52%) | 0.27 |
| **nEV biomarkers, Mean (SD)** |  |  |  |
| α-Synuclein | 0.42 (0.09) | 0.43 (0.07) | 0.44 |
| **Columbia University postmortem assessment (Lewy body pathology)** |  |  |  |
| **Characteristic** | **LBP negative (n=82)** | **LBP positive (n=21)** | **P-value** |
| Women % | 55 (67%) | 14 (67%) | 0.52 |
| APOE ε4% | 30% | 30% | 0.98 |
| Age, Mean (SD) | 85.64 (7.08) | 87.18 (5.68) | 0.11 |
| AD dementia (%) | 42 (51%) | 14 (67%) | 0.22 |
| **nEV biomarkers, Mean (SD)** |  |  |  |
| α-Synuclein | 0.42 (0.19) | 0.54 (0.14) | 0.02 |

**Bio-Hermes**
| Characteristic | Cognitively normal (n=604) | AD dementia (n=307) | P-value |
| --- | --- | --- | --- |
| Women % | 375 (62%) | 150 (49%) | <0.001 |
| APOE ε4% | 34% | 45% | 0.003 |
| Age, Mean (SD) | 70.84 (6.59) | 74.35 (6.24) | <0.001 |
| Amyloid PET positive (%) | 152 (25%) | 149 (49%) | <0.001 |
| P-tau217 positive (%) | 49 (8%) | 77 (25%) | <0.001 |
| **nEV biomarkers, Mean (SD)** |  |  |  |
| α-Synuclein | 0.52 (0.09) | 0.56 (0.10) | <0.001 |

EFIGA, Estudio Familiar de Influencia Genética en Alzheimer, WHICAP, Washington Heights-Hamilton Heights-Inwood Columbia Aging Project, SD, Standard Deviation; nEV, neuronal extracellular vesicle; CSF, Cerebrospinal fluid; AD, Alzheimer's disease; APOE, Apolipoprotein; PET, Positron Emission Tomography.

The PPMI cohort included 83 individuals, of whom 50.6% were healthy and without dementia or PD and 49.4% with PD. nEV-derived α-synuclein and CD9 levels differed significantly between PD and non-demented individuals (*P*<0.05) (Table 1).

The EFIGA cohort consisted of 54 individuals, including 29 (53.7%) without dementia of which 48% were P-tau181 biomarker positive (preclinical AD), and 25 had a clinical diagnosis of dementia with 48% confirmed P-tau181 supported AD. No significant differences (*P*>0.05) were found in age, sex, *APOE* ε4 carrier frequency between the groups. All EFIGA participants underwent cerebrospinal fluid (CSF) α-synuclein seed amplification assay (SAA) testing. Among individuals with preclinical AD, two (7%) were positive for synucleinopathy, compared with 12 (48%) of individuals with AD, representing a significantly higher synucleinopathy in the AD group (*P* =0.01).

The WHICAP cohort included 262 individuals, consisting of 120 (45.8%) without dementia (controls) and 142 (54.2%) with dementia. Age differed significantly (*P*<0.05) between groups (*P*<0.05), whereas sex, *APOE* ε4 frequency, and P-tau181 positivity did not (*P*>0.05). Postmortem neuropathological assessment was available in 103 WHICAP participants, including 82 (79.6%) without and 21 (20%) with Lewy Body pathology. There were no significant differences (*P>*0.05) were observed in age, sex, *APOE* ε4 frequency between individuals with and without Lewy body pathology. In contrast, nEV-derived α-synuclein levels were significantly (*P*<0.05) higher in individuals with Lewy body pathology. We combined the EFIGA and WHICAP cohorts defining this as the Columbia University (CU) dataset.

The Bio-Hermes cohort included 911 individuals, of whom 604 (66.3%) were controls and 307 (33.7%) had AD based on amyloid PET positivity, plasma P-tau217 positivity, and *APOE* ε4 frequency (Table 1). Sex and age also differed significantly between these two groups (Table 1).

Men exhibited significantly higher nEV-derived α-synuclein levels than women in the CU cohort (mean=0.69 vs. 0.60; P=3.59×10□□), with a consistent but non-significant trend in the same direction observed in the Bio-Hermes cohort (*P*>0.05; Supplementary Figure 1A-B). Within P-tau biomarker-positive subgroups, men similarly trended toward elevated nEV-derived α-synuclein levels across both cohorts, though these differences did not reach statistical significance (*P*>0.05; Supplementary Figure 1C-D).

### Association of nEV α-synuclein level with Parkinson’s disease

Plasma nEV-derived α-synuclein levels were CD9-normalized and log and z-score transformed (Figure 1A) prior to analysis. We evaluated whether peripheral nEV-derived α-synuclein levels were associated with clinically diagnosed PD. Compared with non-demented individuals without PD (mean=0.51, SD=0.24), participants with PD (mean=0.69, SD=0.23) exhibited 35.3% higher levels (*P*=8.66×10^-05^) of nEV-derived α-synuclein levels (Figure 1B, Table 2), supporting a strong association between plasma nEV α-synuclein and Parkinson’s disease.

**Figure 1.**
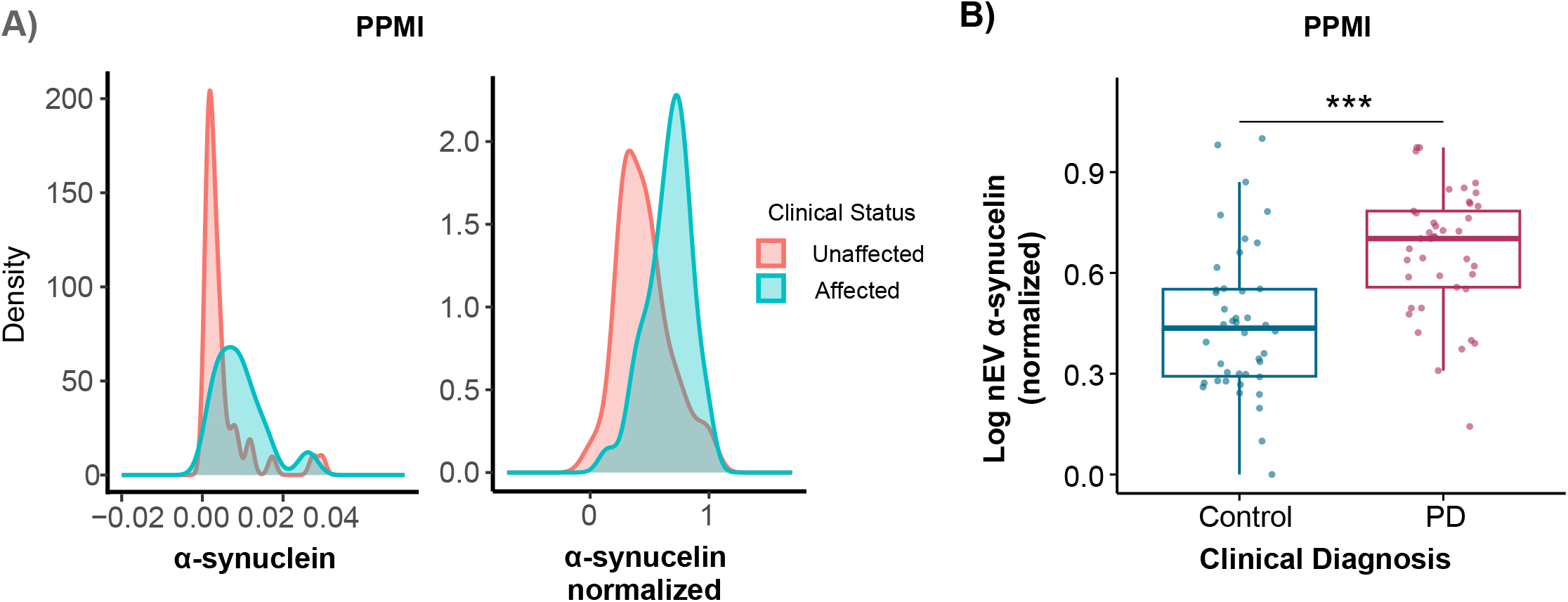
Neuron-derived extracellular vesicle (nEV) α-synuclein levels in Parkinson’s disease (PD). **(A)** Density plots illustrating the distribution of nEV-derived raw α-synuclein levels (left panel) and CD9-normalized, log and z-score transformed α-synuclein measures (right panel) in participants from the Parkinson’s Precision Markers Initiative (PPMI) cohort. **(B)** Box plots demonstrating significantly elevated nEV-derived α-synuclein levels in individuals with PD compared with cognitively normal controls (P < 0.05) from the PPMI cohort. Box plot shows the median and interquartile range. Significance level is provided above comparison group (\**p*<0.05, \*\**p*<0.01, \*\*\**p*<0.001).

**Table 2:** Association of nEV. α**-synuclein levels (outcome) with AD and PD phenotypes (predictor)**

| Predictor | Dataset | N | Mean values in cases | Mean values in controls | Age and sex adjusted ANCOVA model |  | Age and sex adjusted linear regression model |  |  | %diff between cases and controls |
| --- | --- | --- | --- | --- | --- | --- | --- | --- | --- | --- |
|  |  |  |  |  | F-value | P | Beta | T-value | P |  |
| Biomarker Positive AD (Ptau217) | Bio-Hermes | 553 | 0.61 | 0.57 | 17.20 | 3.89E-05 | 0.039 | 3.64 | 2.93E-04 | 7.02 |
| Biomarker Positive AD (Ptau181) | CU | 158 | 0.53 | 0.49 | 2.93 | 0.082 | 0.06 | 2.86 | 4.87E-03 | 8.16 |
| Clinical AD | Bio-Hermes+CU | 1119 | 0.47 | 0.43 | 39.93 | 3.80E-10 | 0.04 | 6.95 | 6.38E-12 | 9.30 |
| SAA | CU | 35 | 0.52 | 0.44 | 1.29 | 0.26 | 0.07 | 1.1 | 0.28 | 18.2 |
| LB pathology | CU | 91 | 0.54 | 0.42 | 5.39 | 0.02 | 0.12 | 2.26 | 0.03 | 28.6 |
| Amyloid PET positivity | Bio-Hermes | 901 | 0.55 | 0.52 | 9.42 | 2.12E-03 | 0.018 | 2.64 | 8.50E-03 | 5.77 |
| Clinical PD | PPMI | 83 | 0.69 | 0.51 | 22.23 | 1.01E-05 | 0.212 | 4.76 | 8.48E-06 | 35.29 |

### Association of nEV α-synuclein levels with biomarker-positive AD and postmortem confirmed Lewy body pathology

First, we evaluated the relationship between nEV-derived α-synuclein levels and AD biomarker positivity across cohorts. Linear regression analyses demonstrated that higher nEV-derived α- synuclein levels were significantly associated with increased P-tau181 levels in the CU cohort (β=0.99, *P*=0.004) (Figure 2A and Supplementary Figure 2A-B) and increased P-tau217 levels in the Bio-Hermes cohort (β=0.30, *P*=0.002) (Figure 2B and Supplementary Figure 2C).

**Figure 2.**
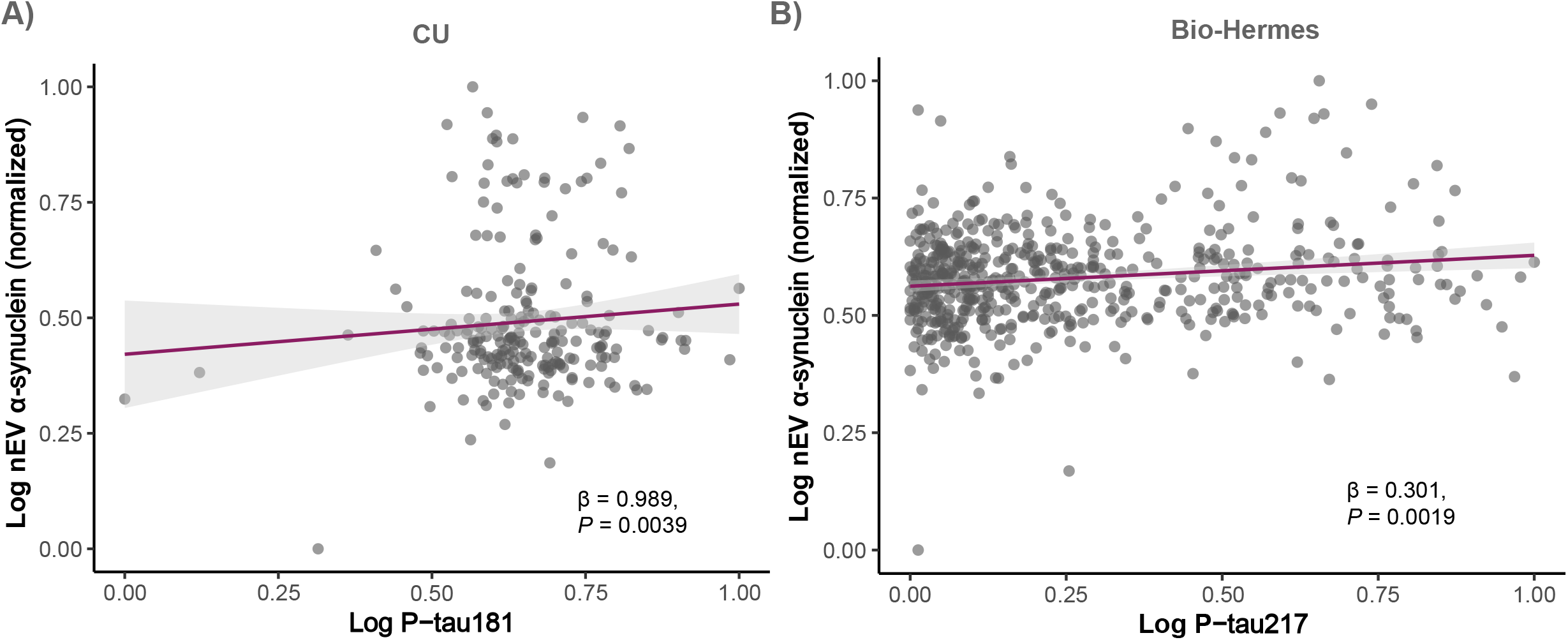
Association between neuron-derived extracellular vesicle (nEV) α-synuclein and P-tau levels. Scatter plots illustrating the linear relationship between nEV-derived α-synuclein levels and P-tau181 in the Columbia University (CU) cohort **(A)**, and between nEV-derived α- synuclein levels and P-tau217 in the Bio-Hermes cohort **(B)**. All nEV-derived α-synuclein measures were CD9-normalized, log-transformed, and z-score standardized. P-tau181 and P- tau217 measures were also log-transformed and z-score standardized.

In the CU cohort, comparing P-tau181-negative individuals with P-tau181-positive individuals showed an 8.2% increase in mean levels of nEV-derived α-synuclein (P-tau181-negative mean=0.49, SD=0.16 vs P-tau positive mean=0.53, SD=0.16, *P*=4.87E-03) (Figure 3A, Table 2). Similar results were observed in the Bio-Hermes cohort. Compared with biomarker-negative individuals, P-tau217 positive individuals demonstrated significantly higher 7.02% increase in nEV-derived α-synuclein levels (P-tau217 negative mean=0.57, SD=0.09 vs P-tau217 positive mean=0.61, SD=0.11, *P*=2.93E-04) (Figure 3A). Collectively, these findings suggest that P-tau-positive individuals exhibit group-wise approximately 7% higher nEV-derived α-synuclein levels than P-tau-negative individuals.

**Figure 3.**
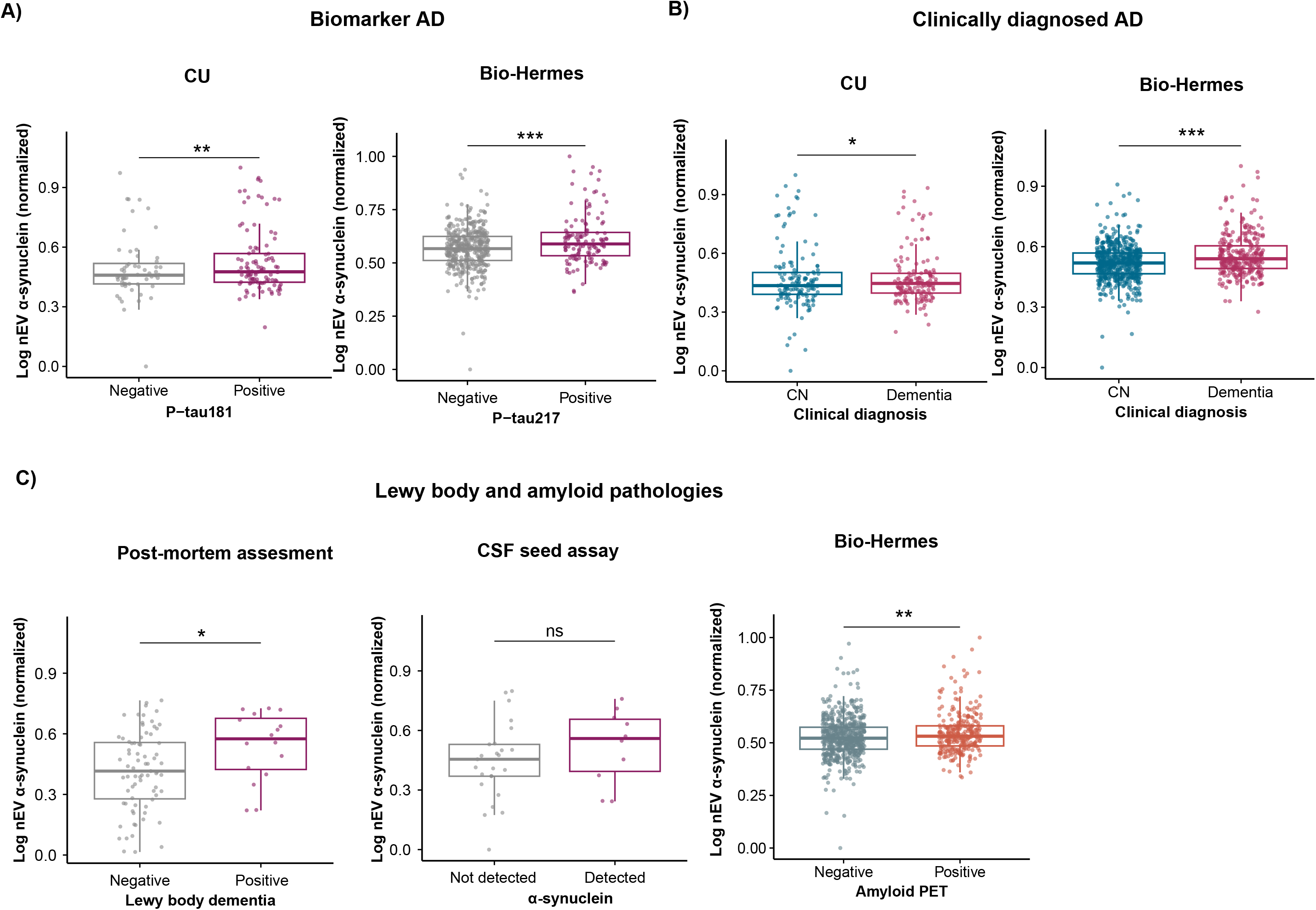
Neuron-derived extracellular vesicle (nEV) α-synuclein levels in pathologically confirmed and clinically characterized cohorts. Box plots illustrating plasma nEV-derived α- synuclein levels across clinically and pathologically defined groups. **(A)** Elevated nEV-derived α-synuclein levels in plasma P-tau181-positive individuals from the Columbia University (CU) cohort (left panel) and in plasma P-tau217-positive individuals from the Bio-Hermes cohort (right panel). **(B)** Elevated nEV-derived α-synuclein levels were observed in individuals with postmortem-confirmed Lewy body pathology compared with controls (left panel). Elevated nEV- derived α-synuclein levels were also observed in cerebrospinal fluid (CSF) α-synuclein seed amplification assay-positive individuals compared with controls (middle panel). Finally, elevated nEV-derived α-synuclein levels were observed in amyloid PET-positive individuals from the Bio-Hermes cohort (right panel). **(C)** Elevated nEV-derived α-synuclein levels in clinically diagnosed dementia patients compared with cognitively normal individuals in the CU (left panel) and Bio-Hermes (right panel) cohorts. All nEV-derived α-synuclein measures were CD9- normalized, log-transformed, and z-score standardized. Box plots represent the median and interquartile range (IQR). Statistical significance is indicated above each comparison (*P* < 0.05, *P* < 0.01, *P* < 0.001).

We tested whether nEV-derived α-synuclein levels were elevated in individuals diagnosed with clinical dementia (with or without AD biomarker support). Individuals with a clinical diagnosis of dementia demonstrated a 9.3% increase in nEV-derived α-synuclein levels compared to healthy individuals (mean=0.47, SD=0.13 vs mean=0.43, SD=0.16, *P*=6.38E-12; Figure 3B). In addition, nEV-derived α-synuclein levels were consistently elevated among mild cognitively impaired (mean=0.55, SD=0.10 vs mean=0.52, SD=0.09, *P*=2.97E-05) and AD dementia (mean=0.56, SD=0.11 vs mean=0.52, SD=0.09, *P*=5.39E-05) in the Bio-Hermes cohort (Supplementary Figure 3). Among WHICAP participants with MRI imaging available, **w**e evaluated whether plasma nEV-derived α-synuclein levels were associated with hippocampal volume. Among individuals with AD dementia, smaller hippocampal volume was correlated (*r*=- 0.23; *P*=0.04) with increased nEV-derived α-synuclein levels, indicating an inverse relationship between hippocampal integrity and peripheral α-synuclein burden (Supplementary Figure 4). These findings suggest that elevated nEV-derived α-synuclein levels are associated with hippocampal atrophy particularly within AD dementia.

To determine whether peripheral nEV-derived α-synuclein reflected underlying Lewy body pathology, we observed increased α-synuclein levels (*P*=0.02) among those with Lewy bodies at postmortem examination (Lewy body negative mean=0.42, SD=0.19 vs Lewy body positive mean=0.54, SD=0.17, *P*=0.02) representing 28.6% increase (Figure 3C, Table 2). These findings validate the association between circulating nEV α-synuclein and synucleinopathy. Consistent with this observation, CSF α-synuclein SAA positive individuals had 18.2% higher nEV-derived α-synuclein levels than individuals with negative α-synuclein seed amplification (SAA positive mean=0.52, SD=0.26 vs SAA negative mean=0.46, SD=0.18). However, due to the small sample size, this association did not reach statistical significance (*P*=0.15) (Figure 3C, Table 2).

We further assessed whether nEV-derived α-synuclein levels were associated with antemortem cerebral amyloidosis as measured by amyloid-PET. In the Bio-Hermes cohort, amyloid PET positive individuals had higher nEV-derived α-synuclein levels compared with amyloid PET negative individuals (mean=0.55, SD=0.10 vs amyloid PET negative mean=0.52, SD=0.09, *P*=8.5E-03; Figure 3C, Table 2).

To further characterize the relationship between AD biomarker status and synucleinopathy, participants we stratified individuals into biomarker-negative controls (BM-CTL), biomarker- positive preclinical individuals (BM+Preclin), biomarker-negative dementia (BM-Dem), and biomarker-positive AD (BM+AD) groups. In the CU cohort, BM+Preclin (mean=0.50, SD=0.14 vs mean=0.48, SD=0.13, *P*=0.03) and BM+AD (mean=0.66, SD=0.19 vs mean=0.48, SD=0.13, *P*=0.006) individuals exhibited significantly higher nEV-derived α-synuclein levels compared with BM-CTL participants (Supplementary Figure 5A). Similarly, in the larger Bio-Hermes cohort, BM+AD (mean=0.48, SD=0.12 vs mean=0.43, SD=0.12, *P*=0.01) and BM-Dem (mean=0.49, SD=0.14 vs mean=0.43, SD=0.12, *P*=6.21E-05) groups showed significantly elevated nEV-derived α-synuclein levels (Supplementary Figure 5B). Collectively, these findings indicate that biomarker positive AD and other dementias are associated with increased peripheral nEV-derived α-synuclein levels, supporting increased occurrence of synuclein co-pathology in biomarker-positive AD and in other dementias.

To support clinical utility of nEV-derived α-synuclein as a biomarker, we estimated an optimal positivity cut-score using the PPMI cohort and WHICAP postmortem cohort with characterized LB pathology. Log-transformed, normalized α-synuclein levels showed clear separation between PD and control distributions, yielding an optimal positivity threshold of z-score > 0.555 (Supplementary Figure 6), and an AUC of 0.78. Applying this derived cutoff to the rest of the CU dataset, we found 46% of P-tau181 positive individuals were also positive for α-synuclein pathology which is consistent with previous AD neuropathology studies^25^. Similarly, 64.7% of postmortem-confirmed Lewy body pathology cases and 66.7% of CSF α-synuclein SAA positive individuals were classified as α-synuclein-positive antemortem.

## Discussion

In this multicohort study, we demonstrated that plasma nEV-derived α-synuclein levels were consistently elevated in PD, biomarker-positive AD, and other dementias, and in individuals with postmortem-confirmed Lewy body pathology. These findings, replicated across multiple independent cohorts, position nEV-derived α-synuclein as a compelling antemortem biomarker of synuclein in PD and co-pathology in AD and related dementias, one that is not achievable with conventional plasma assays.

The original challenge in neurodegeneration biomarker research was insufficient utility of measuring brain protein levels in blood. Conventional plasma biomarkers for some proteins do provide a direct assessment of brain protein levels, but not for others. Specifically, measurements of brain α-synuclein in blood have been very challenging. A possible solution is to use nEVs, since they are released from neurons and traverse the blood-brain barrier into systemic circulation, carrying a molecular fingerprint of neuronal biology. By enriching for this neuronal fraction via immunoaffinity capture of GAP43- and NLGN-expressing vesicles, the ExoSORT platform enables selective measurement of brain-derived α-synuclein from a standard blood draw. This distinction is critical- whole-plasma α-synuclein is confounded by abundant erythrocyte expression, whereas the nEV approach filters this non-specific signal, yielding a central nervous system attributable readout.

This biological specificity is substantiated by the validation of elevated nEV α-synuclein against postmortem-confirmed Lewy body pathology, representing one of the few studies to anchor a blood-based α-synuclein assay to neuropathology. Although the CSF seed amplification assay (SAA) comparison did not reach statistical significance due to the small sample size, the consistent direction and size of effect is encouraging and likely reflects the biological concordance. Inverse correlations between nEV α-synuclein and hippocampal volume specifically among individuals with AD suggest that this peripheral measure might reflect ongoing neurodegeneration in the brain, rather than a peripheral epiphenomenon. Critically, unlike CSF collection, plasma nEV isolation is scalable, repeatable, and amenable to population-level deployment.

Lewy body co-pathology is present in 30–60% of neuropathologically confirmed AD^26–28^ and contributes substantially to clinical heterogeneity, yet has limited detectability with current plasma biomarkers, which are specific to amyloid and tau pathology. Similarly, 40% of individuals with autopsy confirmed PD, have AD as a co-pathology at postmortem examination^29^. Recently developed plasma biomarker panels such as the NUcleic Acid-Linked Immuno-Sandwich Assay (NULISA) central nervous system (CNS) panel show limited to poor detection of synuclein pathology differences between AD patients and healthy controls from plasma^30^. The present findings suggest nEV-derived α-synuclein could fill this diagnostic void by measuring brain-derived levels. The strong and independently replicated associations with P- tau181 and P-tau217 positivity likely reflect the well-characterized co-pathology of amyloid-β, tau, and α-synuclein in the AD brain. Stratification by biomarker status revealed elevated nEV α- synuclein even in biomarker-positive preclinical individuals, suggesting that neuronal synuclein dysregulation might begin early in the AD pathological cascade, an observation with implications for clinical trial enrichment strategies.

Analysis of pathological measures across cohorts, including CSF SAA positivity, postmortem Lewy body pathology, and amyloid PET, demonstrated a consistent and significant overall association with low heterogeneity. The remarkably strong PD association in the PPMI cohort, combined with the AD dementia meta-analysis, demonstrates cross-diagnostic utility spanning the full spectrum of synucleinopathy. This cross-diagnostic validity is clinically meaningful, as Lewy body co-pathology in AD directly affects treatment response, prognosis, and eligibility for α-synuclein-targeting therapies. The nEV platform’s multiplexing potential, enabling simultaneous profiling of α-synuclein alongside tau, amyloid, and synaptic proteins from the same sample further positions it as a candidate component of future multi-marker panels for comprehensive co-pathology characterization. Combining nEVs with multiplexing methodologies like the NULISA panel can further increase biomarker discovery capabilities.

Key limitations include the modest postmortem validation sample (n=31 Lewy Body pathology-positive), the small sample size for the CSF SAA comparison, and the cross-sectional design that precludes examination of longitudinal trajectories. The specificity of neuronal EV enrichment relative to other CNS or peripheral cell types warrants further characterization using single-EV proteomics. Sex differences in nEV α-synuclein, consistently higher in men across cohorts, represent a hypothesis-generating finding that merits dedicated investigation in larger, sex-stratified samples.

Taken together, we found plasma nEVs provide a unique and direct antemortem window into brain α-synuclein levels from a minimally invasive blood draw, a capability not afforded by conventional plasma biomarkers. Validated against postmortem pathology and replicated across multiple cohorts and diagnostic entities, nEV-derived α-synuclein represents a promising biomarker for detecting Lewy body co-pathology in AD and related dementias, with direct applications in diagnosis, patient stratification, and precision therapeutic trial design.

## Methods

### Study participante

#### Estudio Familiar de Influencia Genética en Alzheimer

This study included 54 participants from the Estudio Familiar de Influencia Genética en Alzheimer (EFIGA) cohort, comprising individuals of Caribbean Hispanic ancestry recruited from the Dominican Republic and New York. Detailed descriptions of the study design, participant enrollment, clinical assessments, and standardized medical and neurological examinations have been reported previously ^31^.

Clinical severity was assessed using the Clinical Dementia Rating (CDR) scale ^32^. AD diagnoses were established according to the National Institute of Neurological and Communicative Disorders and Stroke-Alzheimer’s Disease and Related Disorders Association (NINCDS-ADRDA) criteria ^33–35^. Additional details regarding the EFIGA cohort have been described elsewhere ^31^.

#### CSF collection

CSF was collected by aseptic lumbar puncture, centrifuged at 2,000 × g for 10 minutes at 4°C, aliquoted into low-binding polypropylene tubes, and stored at −80°C until analysis with no more than two freeze-thaw cycles.

#### α-Synuclein Seed Amplification Assay (SAA))

CSF α-synuclein seed amplification assay (SAA) was performed by Amprion in their CLIA validated laboratory using the SAAmplify assay. Samples were analyzed in triplicate (40 uL per well) in 96-well plates in 200 μL reaction volume containing recombinant monomeric human α- synuclein, and thioflavine T, using intermittent shaking and bottom fluorescence measurement of ThT fluorescence (excitation 440 nm, emission 490 nm). All assays were performed blinded to clinical and biomarker data.

#### Washington Heights/Inwood Columbia Aging Project

This study included 262 participants from the Washington Heights-Hamilton Heights-Inwood Columbia Aging Project (WHICAP), a prospective, community-based cohort study designed to investigate aging and dementia among older adults residing in northern Manhattan, New York. WHICAP enrolled participants aged 65 years and older through three recruitment waves initiated in 1992, 1999, and 2009 using comparable study protocols and assessment procedures. At study enrollment, participants underwent a structured interview assessing demographic characteristics, medical history, general health, and functional status. Comprehensive clinical evaluations were subsequently performed, including standardized medical, neurological, and psychiatric assessments, as well as detailed neuropsychological testing. Clinical diagnoses were established using consensus-based evaluations integrating medical history, neurological examination, and cognitive performance measures ^36,37^.

All clinical and neuropsychological data were independently reviewed by a multidisciplinary panel comprising two physicians and two neuropsychologists with expertise in dementia diagnosis. Diagnostic evaluations incorporated evidence of cognitive, social, and occupational functional impairment. Dementia diagnoses of all etiologies were determined according to criteria from the Diagnostic and Statistical Manual of Mental Disorders (DSM-IV and DSM- 5)^38,39^. AD diagnoses were established using the National Institute on Aging - Alzheimer’s Association (NIA-AA) criteria for probable or possible AD^40^. Participants who did not meet criteria for dementia were classified as having mild cognitive impairment (MCI)^41^ if they demonstrated impairment in one or more cognitive domains or reported subjective memory complaints while maintaining preserved activities of daily living. Amnestic MCI (aMCI) was defined according to Petersen criteria as previously described ^42–44^.

#### Blood-based biomarkers (WHICAP and EFIGA)

Blood samples were collected by standard venipuncture into dipotassium ethylenediaminetetraacetic acid (K2EDTA) tubes. Plasma was isolated within 2 hours of blood collection by centrifugation at 2,000 × *g* for 15 minutes at 4°C. Following centrifugation, plasma samples were aliquoted into polypropylene tubes, stored at -80°C, and maintained until biomarker analysis. Plasma biomarker measurements were performed using the single-molecule array (Simoa) technology^45^ on the Quanterix HD-X platform (Quanterix, Billerica, MA, USA). All samples were assayed in duplicate using the Quanterix P-tau181 Advantage V2 assay kit (Catalog No. 103714) according to the manufacturer’s instructions ^37^.

#### Lewy Body Pathology (WHICAP)

Lewy body pathology was assessed using α-synuclein immunohistochemistry across brainstem, limbic, and neocortical regions to classify using the McKeith consensus criteria. Any α-synuclein pathology in a case was classified as Lewy body positive.

#### Magnetic Resonance Imaging (WHICAP)

Structural magnetic resonance imaging (MRI) scans for WHICAP participants were acquired using scanners with 1.5T and 3.0T magnetic field strengths. Detailed MRI acquisition and processing protocols have been described previously. Hippocampal volumes were quantified using FreeSurfer according to previously established procedures^46^.

#### Bio-Hermes

This study included 911 participants from the Bio-Hermes study. Participants were recruited between April 2021 and November 2022 across 17 research sites from community-based populations. These sites had prior experience conducting clinical trials evaluating investigational therapies for AD and were experienced in protocols incorporating brain amyloid positivity for study enrollment. Participants were categorized as CN, MCI, or mild AD ^47^.

Eligible participants were aged 60-85 years and fluent in the language used for cognitive testing at the participating site. English-language assessments were approved at all sites, whereas Spanish-language assessments were available at eight sites. Participants completed the Mini-Mental State Examination (MMSE) and Geriatric Depression Scale (GDS)^48^. Additional eligibility criteria included no known negative brain amyloid positron emission tomography (PET) scan within the preceding 12 months, no history of cancer within the past 5 years (excluding melanoma in situ or prostate carcinoma in situ), and no history of seizures or stroke within the previous 12 months. Participants with unstable medical conditions likely to affect cognitive performance or with other conditions incompatible with participation in therapeutic trials were excluded ^47^.

CN participants were defined as having no evidence of memory impairment or subjective cognitive complaints, MMSE scores ranging from 26 to 30, and Rey Auditory Verbal Learning Test (RAVLT) delayed recall scores^49^ within the normal range (>1.5 standard deviations above age and race-adjusted means)^50^. Preserved functional abilities were confirmed using the Functional Activities Questionnaire (FAQ)^51^. MCI participants were classified according to National Institute on Aging-Alzheimer’s Association (NIA-AA) criteria ^52^, with MMSE scores ranging from 24 to 30 and RAVLT delayed recall scores at least 1 standard deviation below age- and race-adjusted means. Participants additionally exhibited minimal to mild cognitive impairment with preserved activities of daily living based on investigator assessment. Mild AD participants met NIA-AA criteria^52^ for probable AD and had MMSE scores ranging from 20 to 24, although investigator discretion permitted inclusion of selected individuals with MMSE scores as low as 17. Participants also demonstrated RAVLT delayed recall scores at least 1 standard deviation below age and race-adjusted means together with evidence of functional decline and impaired independence in daily activities. Additional details regarding the Bio-Hermes cohort have been described previously ^47^.

#### Blood based biomarker (Bio-Hermes)

Blood samples were collected into five K2 EDTA tubes and centrifuged at 1,500 (±100) × g for 15 minutes. Plasma samples were processed and frozen at -80°C within 4 hours of blood collection, with total blood volume limited to ≤50 mL per study visit. Samples remained stored at -80°C prior to shipment. Temporary storage at -20°C was permitted for a maximum of 48 hours before transfer to -80°C storage. Plasma samples were subsequently shipped to the Eli Lilly and Company Clinical Diagnostics Laboratory (Indianapolis, IN, USA) for measurement of plasma P-tau217 ^47^.

#### Brain amyloid PET imaging (Bio-Hermes)

Amyloid PET imaging was performed at designated imaging facilities associated with each participating site. Participants received an intravenous injection of florbetapir F-18 (Amyvid 18F-AV-45) according to standardized administration procedures, followed by PET acquisition approximately 20 minutes after tracer administration. PET images were uploaded and maintained within a centralized electronic imaging repository. Standardized uptake value ratio (SUVR) measurements were converted to the Centiloid scale to harmonize amyloid burden estimates across sites and imaging protocols. Detailed PET acquisition and image-processing procedures have been described previously ^47^.

#### Parkinson Progression Marker Initiative (PPMI)

This study included 83 participants from the Parkinson’s Progression Markers Initiative (PPMI) cohort with available plasma nEV α-synuclein measurements and clinical diagnostic information. PPMI is an ongoing, multicenter observational study designed to identify biomarkers of Parkinson’s disease (PD) progression, with detailed study design and recruitment procedures described previously. Participants with PD were required to have a diagnosis of early, untreated PD with symptom onset within 2 years of enrollment. Clinical eligibility criteria included the presence of resting tremor, asymmetric resting tremor, asymmetric bradykinesia, or a combination of bradykinesia and rigidity. All participants with PD underwent dopamine transporter (DAT) imaging, and evidence of DAT deficit was required to confirm PD eligibility. Healthy control participants were required to have no history of neurological disorders, a Montreal Cognitive Assessment (MoCA) score greater than 26, and no first-degree family history of PD. Additional details regarding participant recruitment, eligibility criteria, and clinical assessments have been described previously ^24^.

#### Neuron-Derived Extracellular Vesicles (nEVs) enrichment

nEVs were enriched from human plasma using ExoSORT platform, an immunoaffinity-based workflow designed to predominantly capture extracellular vesicles of neuronal origin. Blood samples were collected in K2EDTA vials, processed for plasma preparation using standard procedure and underwent no more than 2 freeze-thaw cycles prior to EV isolation.

ExoSORT was performed as previously described (Eitan, Thornton-Wells et al. 2023, Lucafo, Bidoli et al. 2025, Evers, Watson et al. 2026), with modifications for the first automated version. Plasma aliquots (400 µL) were transferred to a deep-well 96-well plate containing ExoSORT plasma diluent (200 µL), mixed, and incubated with total EV (TEV) isolation reagent (250 µL/sample). TEVs were precipitated by centrifugation and resuspended in ExoSORT Binding Buffer (500 µL).

For automated neuron-derived EV (nEVs) capture and washing, magnetic capture beads coupled to antibodies against GAP43 and NLGN were added, and samples processed on a KingFisher instrument (Thermo Fisher Scientific). After four washes, bead–antibody complexes were transferred to ExoSORT elution buffer (70 µL per sample), incubated 5 min, and beads discarded.

Eluates were collected into a storage plate; 30 µL aliquots were used for QC (LuminEV), and the remainder lysed with ExoSORT lysis buffer (150 µL) and stored at −80°C.

#### LuminEV analysis of NDE surface proteins

LuminEV was performed as described (Tordoff, Allen et al. 2024). Briefly, MagPlex microspheres (Diasorin) were functionalized with antibodies against CD9, CD63, CD81, and Grin2A; IgG controls were included.

NDE eluates were diluted 1:3, plated in duplicate (50 µL/well), and incubated overnight at 4°C with capture beads (5 × 10 □ beads/mL per set). After washing, detection was performed using a biotinylated pan-tetraspanin antibody cocktail (CD9, CD63, CD81), followed by streptavidin-PE. Beads were washed, resuspended in sheath fluid, and analyzed on a Luminex xMAP Intelliflex system (≥50 beads/analyte). Results are reported as median fluorescence intensity (MFI), corrected for background.

#### Alpha-synuclein nEV measurements

Oligomeric α-synuclein (α-Syn) was measured using an in-house Luminex-based sandwich immunoassay. MagPlex beads were coupled with anti-α-Syn aggregate antibody (MJFR-14-6-4- 2; Abcam) or isotype control antibodies.

Standards (7.6–5000 pg/mL) and diluted NDE samples were incubated with capture beads overnight at 4°C. After washing, biotinylated detection antibody (4B12, BioLegend) and streptavidin-PE were applied sequentially. Beads were resuspended in sheath fluid and analyzed on the Intelliflex system.

#### Statistical Analysis

Participants from the PPMI, Columbia University (CU) EFIGA, CU WHICAP, and Bio-Hermes cohorts were categorized into unaffected and affected groups based on their clinical diagnoses. In the CU EFIGA and CU WHICAP cohorts, biomarker-positive individuals were defined using a plasma P-tau181 cutoff of 2.648 pg/mL ^53^. In the Bio-Hermes cohort, biomarker-positive individuals were defined using a plasma P-tau217 cutoff of 0.39 pg/mL^37^. Amyloid PET positivity was defined as a Centiloid value >30 ^54^.

Differences in demographic, clinical, biomarker, and pathological characteristics between groups were assessed using the χ² test for categorical variables and the Kruskal-Wallis test for continuous variables in the R software version 4.5.1 (https://www.r-project.org/). Statistical significance was defined as a two-sided *P*<0.05.

The distribution of nEV-derived α-synuclein levels was visualized using the ‘ggplot2’ package^55^ in R software. Raw nEV-derived α-synuclein measurements were normalized to CD9, log and z-score transformed, and subsequently scaled using min-max normalization ^56^. Associations between nEV-derived α-synuclein levels and P-tau measures were performed by linear regression model adjusting for age and sex as covariates in the CU cohort, whereas models in the Bio-Hermes cohort were additionally adjusted for assay batch and recruitment site.

Associations between nEV-derived α-synuclein levels and clinical, biomarker positivity status, and pathological outcomes were evaluated using logistic regression models. Analyses of Parkinson’s disease status in the PPMI cohort were adjusted for sex. Associations with P-tau181 biomarker status, postmortem-confirmed Lewy body pathology, and CSF α-synuclein seed amplification assay status in the CU cohorts were evaluated using logistic regression models adjusted for age and sex. For analysis involving postmortem-confirmed Lewy body pathology, extreme outliers were excluded if nEV-derived α-synuclein levels exceeded 1.5 times the interquartile range above the third quartile (Q3) or below the first quartile (Q1). Associations with amyloid PET positivity, P-tau217 biomarker status, and clinical diagnosis in the Bio-Hermes cohort were evaluated using logistic regression models adjusted for age, sex, assay batch, and recruitment site. Odds ratios (ORs) and corresponding 95% confidence intervals (CIs) were reported for all regression analyses.

To evaluate differences in nEV-derived α-synuclein levels across clinical, biomarker, and pathological groups, age and sex-adjusted analyses of covariance (ANCOVA) and linear regression models were performed with nEV-derived α-synuclein levels as the dependent variable. ANCOVA models were used to compare adjusted mean nEV-derived α-synuclein levels between groups, and statistical significance was assessed using the F statistic and corresponding two-sided *P* values. Linear regression models were additionally used to estimate the magnitude and direction of associations, and standardized regression coefficients (β), *t* statistics, and *P* values were reported. Analyses involving P-tau181 biomarker status, CSF α-synuclein seed amplification assay status, and postmortem-confirmed Lewy body pathology in the CU cohorts were adjusted for age and sex, whereas analyses in the Bio-Hermes cohort were additionally adjusted for assay batch and recruitment site.

Associations between hippocampal volume and nEV-derived α-synuclein levels were evaluated separately within non-dementia and AD dementia groups using partial Pearson correlation analyses adjusted for age, sex, intracranial volume, and MRI magnetic field strength. Partial correlations were performed using the ‘ppcor’ package^57^, and scatter plots were generated using the ‘ggpubr’ package^58^ in R software.

## Data Availability

All intermediate and final results are included in the main text or the supplementary materials of the manuscript. The nEV biomarkers, autopsy, α-synuclein SAA and phenotype data from the EFIGA and WHICAP cohorts are available upon request here (https://cumc.co1.qualtrics.com/jfe/form/SV_dmck0uV3A91pmzb
https://cumc.co1.qualtrics.com/jfe/form/SV_6x5rRy14B6vpoqN).
The Bio-Hermes data was accessed through the Alzheimer's disease data initiative database platform (https://www.alzheimersdata.org/). The PPMI data was accessed through https://www.ppmi-info.org/access-data-specimens/download-data.The nEV biomarker data generated in Bio-Hermes and PPMI cohorts is also available upon request through NeuroDex.

## Data Availability

All intermediate and final results are included in the main text or the supplementary materials of the manuscript. The nEV biomarkers, autopsy, α-synuclein SAA and phenotype data from the EFIGA and WHICAP cohorts are available upon request here (https://cumc.co1.qualtrics.com/jfe/form/SV_dmck0uV3A91pmzb https://cumc.co1.qualtrics.com/jfe/form/SV_6x5rRy14B6vpoqN).

The Bio-Hermes data was accessed through the Alzheimer’s disease data initiative database platform (https://www.alzheimersdata.org/). The PPMI data was accessed through https://www.ppmi-info.org/access-data-specimens/download-data.The nEV biomarker data generated in Bio-Hermes and PPMI cohorts is also available upon request through NeuroDex.

## Code Availability

We used standard packages implemented in R for association, AUC and principal component analyses. R-code used for the analyses in this study is available from the corresponding authors upon request. Interested researchers are encouraged to contact the corresponding authors with a brief description of the intended use. Access will be provided for noncommercial purposes and solely for reproducing the results presented in this Article.

## Acknowledgements

We thank Dr. Jennifer Lamoureux at Amprion Diagnostics for leading the α-synuclein seed assay experiments in the CU cohort.

Data collection for this project was supported by the Genetic Studies of Alzheimer’s disease in Caribbean Hispanics (EFIGA) funded by the National Institute on Aging (NIA) and by the National Institutes of Health (NIH) (R56AG063908, R01AG067501 and RF1AG015473). We acknowledge the services of CEDIMAT for collaborating with sample collection and processing in the EFIGA cohort. WHICAP is funded by the National Institute on Aging (NIA) and by the National Institutes of Health (NIH) (WHICAP, R01AG072474, R01AG066107).

Funding was provided by the Alzheimer’s Drug Discovery Foundation Diagnostics Accelerator and the Michael J. Fox Foundation (MJFF) to NeuroDex, which also provided access to the PPMI samples. This work was also supported by NINDS grant (R44NS130864)

## Author contributions

Conceptualization: RPM, EE, BNV

Methodology: TIG, RPM, OV, EE, BNV

Participant enrolment and Sample Collection: DRD, LSH, RPM, DB, LA

Data Generation and analysis and interpretation: TIG, YG, OV, AT, LSH, RPM, EE, RPM, BNV

Funding acquisition: RPM, BNV

Project administration: DRD, EE, RPM

Supervision: BNV, EE, RPM

Writing – original draft: TIG, EE, RPM, BNV

Writing – review & editing: All authors

