## Supplementary Figure 1-6 for "Antemortem α-Synuclein levels from neuron-derived extracellular vesicles identify Lewy Body pathology in Parkinson’s Disease and Alzheimer Disease"

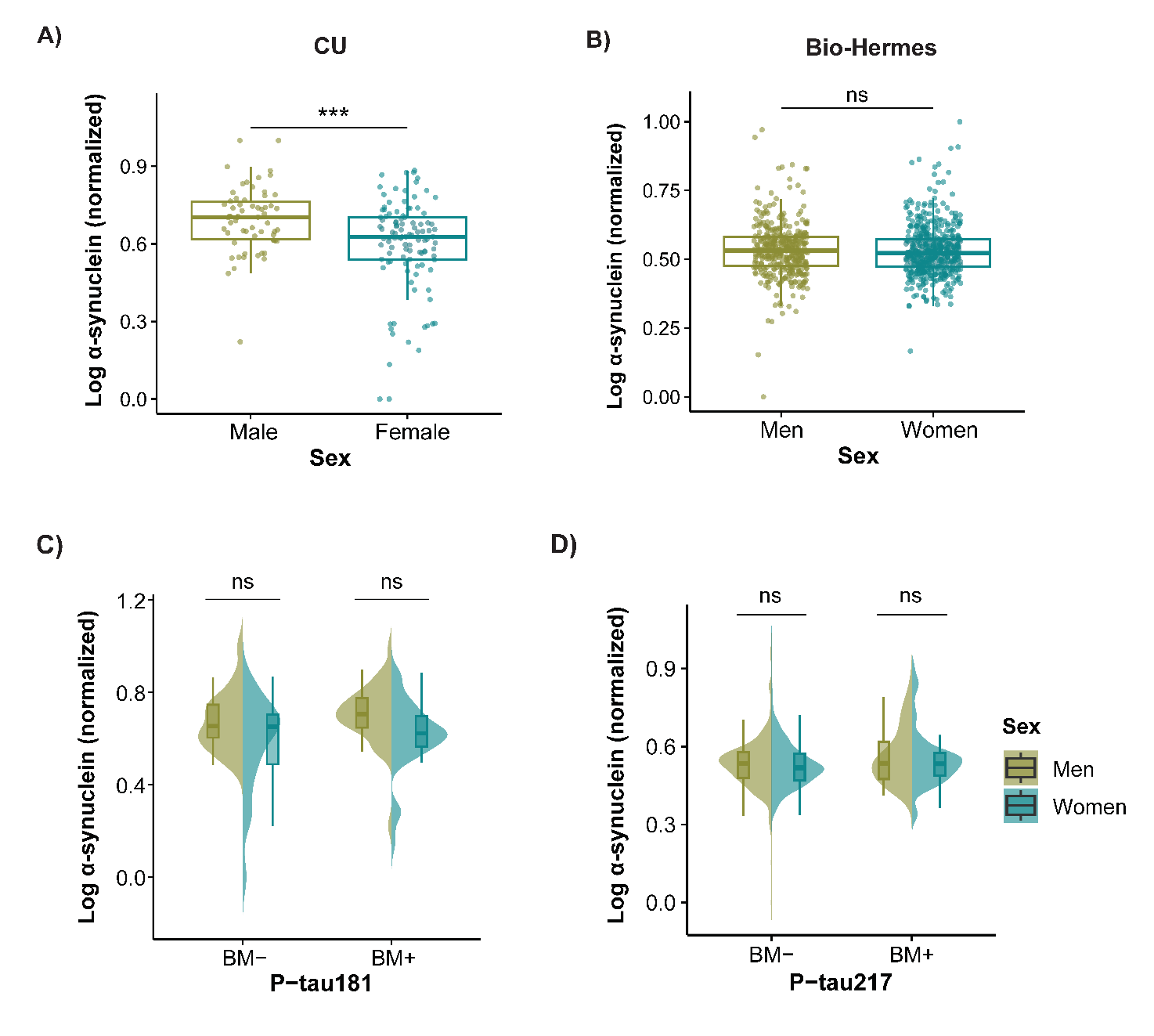

**Supplementary Figure 1. Sex differences in neuron-derived extracellular vesicle (nEV) α-synuclein levels across cohorts. (A-B)** Box plots comparing nEV-derived α-synuclein levels between men and women in **(A)** the Columbia University (CU) cohort, and **(B)** the Bio-Hermes cohort. **(C-D)** Split violin plots showing nEV-derived α-synuclein levels compared between men and women stratified by biomarker status, including P-tau181 negative and P-tau181 positive individuals in **(C)** the CU cohort and **(D)** the Bio-Hermes cohort. All nEV-derived α-synuclein measures were CD9-normalized and log and z-score transformed. Statistical significance is indicated above comparison groups (*P*<0.05, ** *P*<0.01, *** *P*<0.001, ns = not significant). Men are shown in green and women in aqua blue.

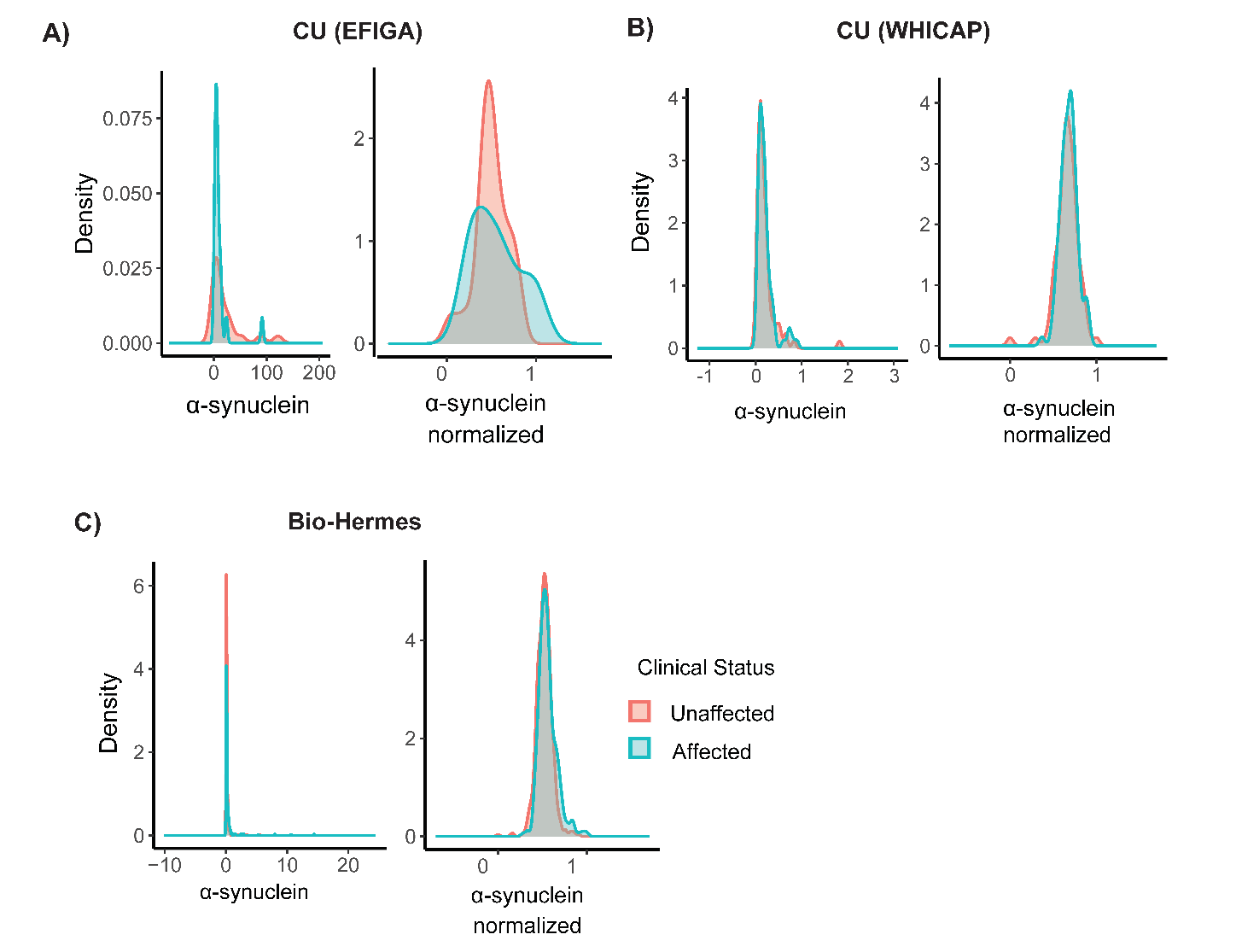

**Supplementary Figure 2. Distribution of neuron-derived extracellular vesicle (nEV) α-synuclein levels across cohorts.** Density plots illustrating the distribution of nEV-derived raw α-synuclein levels (left panels) and CD9-normalized, log and z-score transformed α-synuclein measures (right panels) in participants from **(A)** the Columbia University *Estudio Familiar de Influencia Genética en Alzheimer* (EFIGA) cohort, **(B)** the Columbia University *Washington Heights-Hamilton Heights-Inwood Columbia Aging Project* (WHICAP) cohort, and **(C)** the Bio-Hermes cohort.

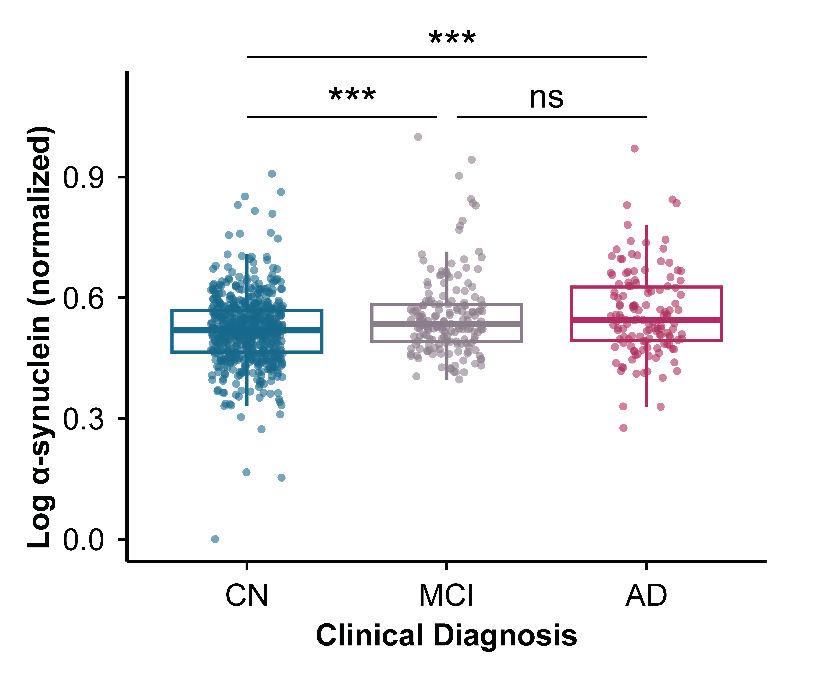

**Supplementary Figure 3. Comparison of neuron-derived extracellular vesicle (nEV) α-synuclein levels across clinically diagnosed groups.** Box plots showing nEV-derived α-synuclein levels across cognitively normal (CN), mild cognitively impaired (MCI) and Alzheimer’s disease (AD) in the Bio-Hermes cohort. All nEV-derived α-synuclein measures were CD9-normalized and log and z-score transformed. Box plots display the median and interquartile range (IQR). Statistical significance is indicated above comparison groups (*P*<0.05, ** *P*<0.01, *** *P*<0.00, ns = not significant).

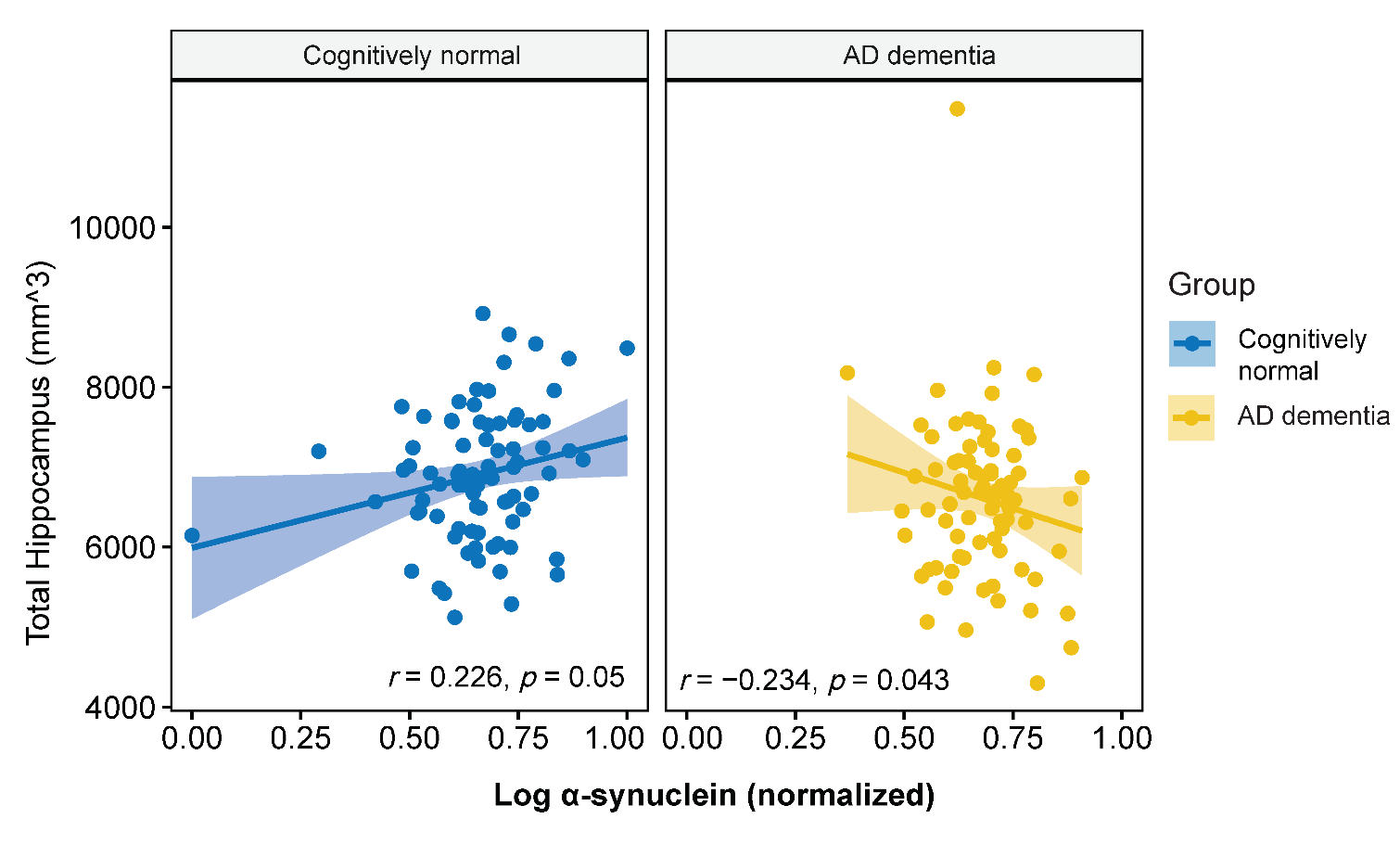

**Supplementary Figure 4. Correlation of nEV-derived α-synuclein levels with hippocampal volume.** Scatter plot shows the correlation between hippocampal volume and nEV-derived α-synuclein levels stratified by clinical diagnosis. The left panel shows cognitively normal individuals, indicated in blue, and the right panel shows individuals with Alzheimer’s disease dementia, indicated in yellow.

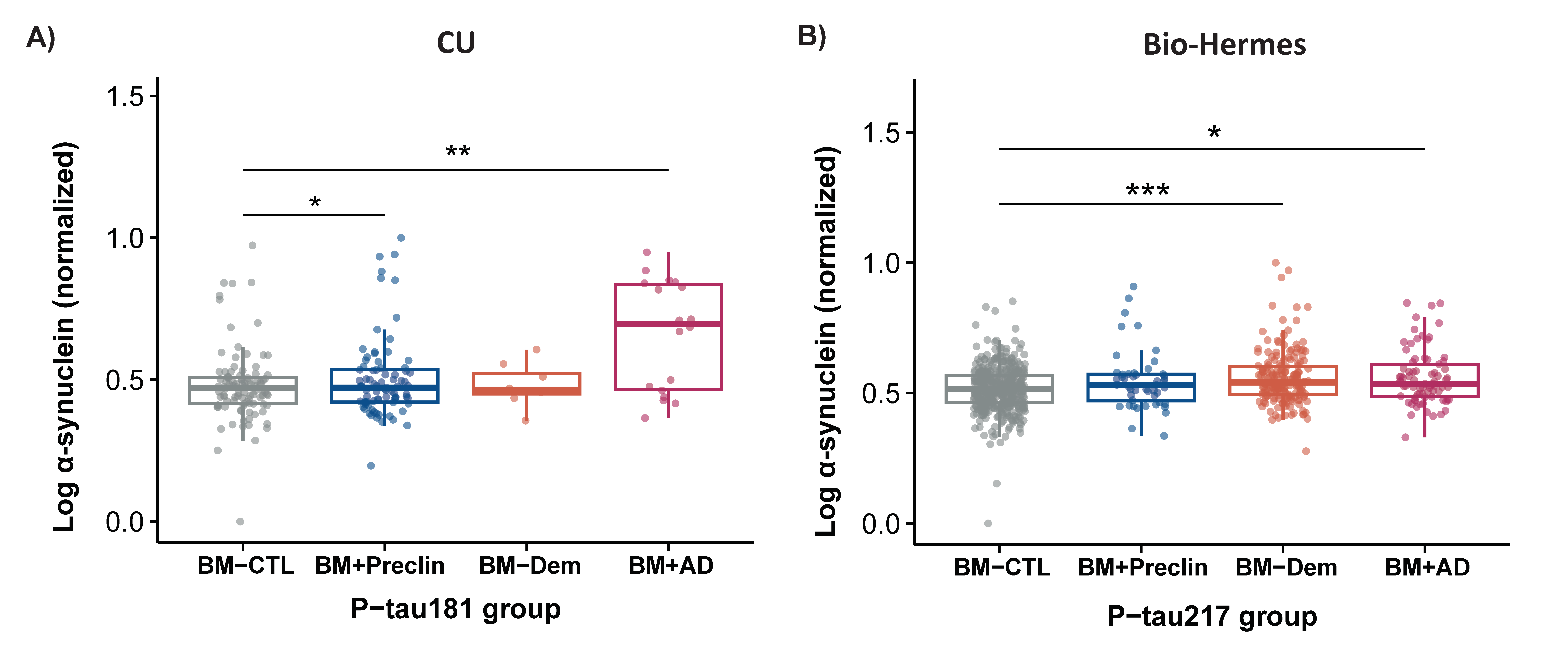

**Supplementary Figure 5. Comparison of neuron-derived extracellular vesicle (nEV) α-synuclein levels across P-tau defined clinical subgroups. (A)** Box plots showing nEV-derived α-synuclein levels across clinical groups stratified by phosphorylated tau at threonine 181 (P-tau181) biomarker status in the Columbia University cohort. **(B)** Box plots showing nEV-derived α-synuclein levels across clinical groups stratified by P-tau217 biomarker status in the Bio-Hermes cohort. Clinical groups were categorized as biomarker-negative controls (BM-CTL), biomarker-positive preclinical individuals (BM+Preclin), biomarker-negative dementia (BM-Dem), and biomarker-positive Alzheimer’s disease (BM+AD). All nEV-derived α-synuclein measures were CD9-normalized and log and z-score transformed. Box plots display the median and interquartile range (IQR). Statistical significance is indicated above comparison groups (*P*<0.05, ** *P*<0.01, *** *P*<0.00, ns = not significant).

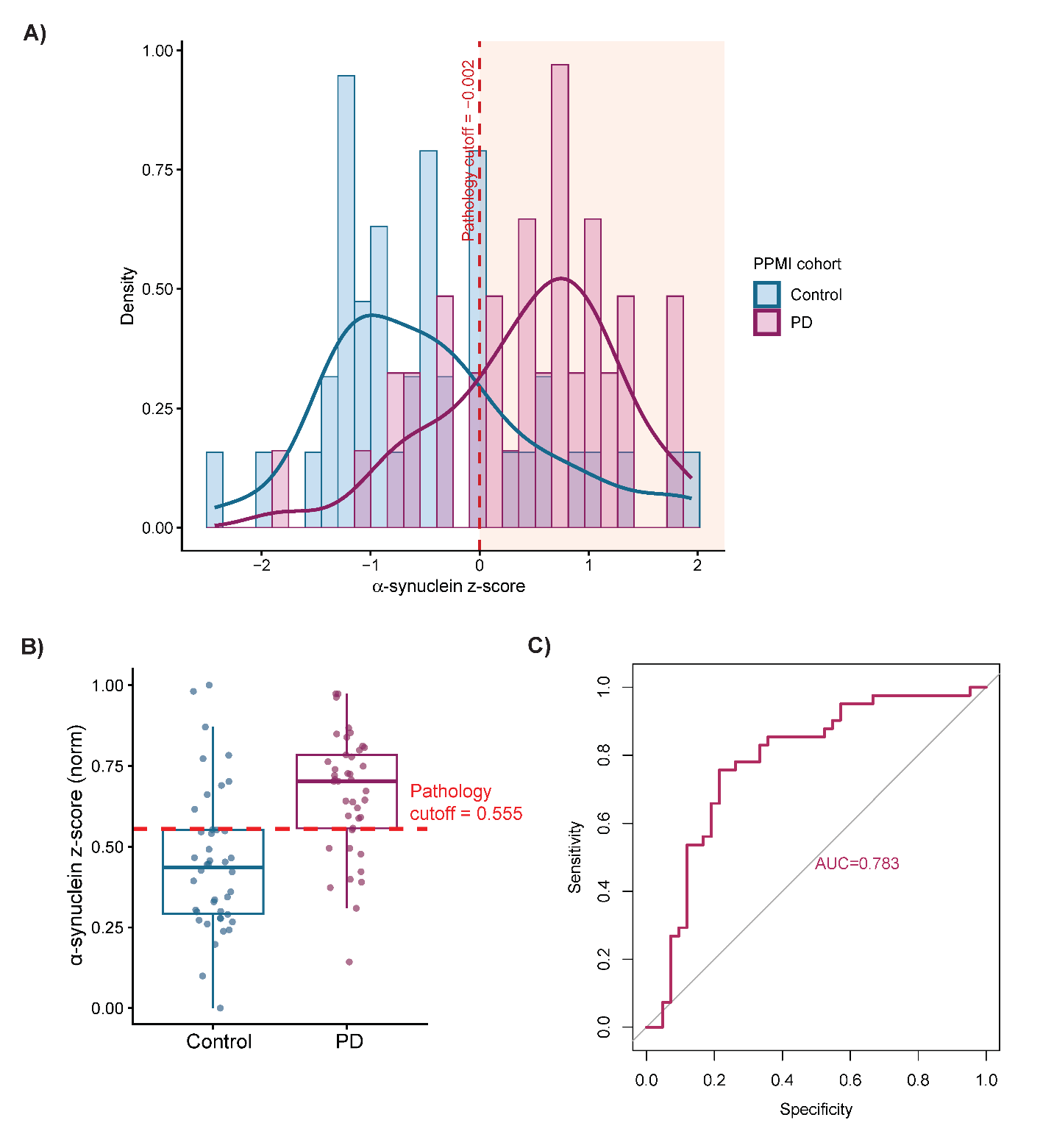

**Supplementary Figure 6. Estimation and visualization of the α-synuclein cut-score. (A)** Distribution of log-transformed, z-score normalized nEV-derived α-synuclein levels in the PPMI cohort. The histogram shows the density distribution of α-synuclein levels in individuals with Parkinson’s disease (PD) in maroon and controls in light blue, together with the optimal α-synuclein positivity cut-score (z-score > -0.002), indicated by the red dashed line. **(B)** The corresponding box plots display the distribution of α-synuclein levels using z-score and min-max normalization. The red dashed line represents the estimated α-synuclein positivity threshold in box plot. **(C)** ROC curves showing the predictive performance of nEV-derived α-synuclein for distinguishing PD (maroon). The corresponding AUC values quantify the overall discriminatory ability of the biomarker.
